# Racialised experience of detention under the Mental Health Act: a photovoice investigation of practice, policy and legislation

**DOI:** 10.1101/2024.09.30.24314284

**Authors:** Kamaldeep Bhui, Roisin Mooney, Doreen Joseph, Rose McCabe, Karen Newbigging, Paul McCrone, Ragu Raghavan, Frank Keating, Nusrat Husain

**Affiliations:** Department of Psychiatry, University of Oxford; Nuffield Departmenwt of Primary Care Health Sciences, University of Oxford; Wadham College, University of Oxford; School of Health Sciences, City University; School of Social Policy, University of Birmingham; Institute for Life Course Development, Greenwich University; Health and Life Sciences, De Montfort University; Dept of Law and Criminology, Royal Holloway University of London; Division of Psychology and Mental Health, University of Manchester & Co-Pact Team

## Abstract

**Background:** The rates of compulsory admission and treatment (CAT) are rising in mental health systems in the UK. These disparities are reported among migrants and black and ethnic minorities in Europe and North America. Lived experience perspectives from marginalised and multiple disadvantaged people are neglected in research yet may offer vital and novel insights into preventive opportunities to reduce coercive care

**Methods:** We conducted a participatory photovoice research process to assemble the life-experiences of people within two years of receiving CAT. We purposively sampled to maximise diversity by age, ethnicity, and different ‘sections’ of the Mental Health Act (England & Wales, 1983, 2007) from 8 health systems in England. The images, captions, and reflective narratives were deepened over 3 workshops before thematic analysis.

**Outcomes:** Forty-eight ethnically diverse people with lived experience of CAT contributed over 500 images and 30 hours of transcribed narratives. Participants lives showed significant complexity in terms of multimorbidity, adverse childhood experiences, and carer roles. The findings suggest insufficient co-ordination to prevent CAT despite early help-seeking, not being taken seriously when seeking help, hostility and dismissive responses from professionals; unnecessary police involvement which was distressing, stigmatising, and risked criminalisation. Participants wanted more advocacy given their vulnerability and inability to process information in crisis, as well as therapeutic and creative activities in inpatient environments. Participants recommended more family and carer involvement, and more appropriate, frequent and personalised information about care options, appeals processes, levels of restriction, and seclusion. A major concern was the lack of highly skilled staff, trauma-informed care, and therapies into the community.

**Interpretation:** We showed epistemic injustice in care processes and recommend better standards for essential skills, prevention, trauma informed care and therapies. Criminalising and coercive responses must also be reduced.

**Funding:** This study/project is funded by the National Institute for Health Research (NIHR) Policy Research Programme [NIHR201704]. The views expressed are those of the author and not necessarily those of the NIHR or the Department of Health and Social Care

## Background

Ethnic minorities and migrants in the UK experience more adverse pathways to mental health care, including higher rates of compulsory admission and treatment (CAT), more contact with criminal justice agencies, and poorer long term outcomes compared with White British people.^1–5^ These data also reflect the picture in North America and Europe.^6–8^ Since 2008, rising rates of detention in England and Wales have cost an extra £6.8 billion.^9^ Therefore reducing detentions might release savings, which could be re-invested to improve community-based support, alternatives to detention, early intervention, and crisis care.^10^ Reducing CAT may also mitigate stigma, feelings of powerlessness and fear of services.^11–14^

Actions to reduce CAT must target causal drivers, however, most explanations focus on individual characteristics rather than care systems.^15^ For example, potential explanations include traumatic psychosocial adversities over the life-course, such as early separation from parents, discrimination, isolation, unemployment, living in poorer places, gender violence, substance use and health risk behaviours, and psychiatric co-morbidities.^16^ ^1^ There may also be ethnicity related variations in help-seeking behaviours, clinical assessment of symptoms and dangerousness; poorer therapeutic alliances may also contribute.^3^ ^17^ ^18^ ^16^ ^6^ ^1^ Some argue that escalating levels of CAT arise due to delays in accessing effective services because of the stigma associated with mental illness as well as fears about harms and coercion.^19^ Yet, some studies show there is no delay in help seeking for first episodes of psychosis among ethnic minorities.^20^ The higher rates of CAT may reflect crises due to the absence of carers or lack of primary care involvement.^21^ In addition, austerity measures reduced service capacity and may undermine the prevention of CAT. ^22^ An Independent Review of the Mental Health Act (MHA England & Wales) recommended more research including the experiences of those subject to CAT to inform actions to reduce CAT.^1^ ^2^

The research aims to:

- Assemble experience data from racially and ethnically diverse people who experienced CAT under the powers of the Mental Health Act.
- Identify pathways to care and influences (biographical, interpersonal, societal, situational) leading to CAT, and related preventive lessons.
- Capture care experiences to inform progressive care, policy and legislation.

## Methods

### Inclusive Research

Ethnic minorities and marginalised groups are under-represented in research.^23–26^ Accessibility and language barriers, fluctuating levels of insight and capacity, and fear of losing freedoms may lead to avoidance of services and research institutions if previous experiences were unsatisfactory and traumatic.^27^ ^28^ People may also struggle to verbalise painful memories. Furthermore, research processes may not support participation of the most marginalised, living with multiple disabilities, frailties, and precarious social situations.^29^ ^30^

Therefore, in this study, we draw on participatory qualitative research methods, which can flexibly explore experiences to discover new perspectives. Such approaches have shown complex interactions between environmental, social, and biological drivers of health inequalities. For example, these methods have explained co-existence of diabetes, depression, and HIV, and why these combinations are more common in certain places and among those facing multiple disadvantage at the intersections of ethnicity and gender.^20^ ^31^ ^32^ ^33^

We take a discovery orientated approach that foregrounds lived experience, as proposed in critical studies of black feminism, race theory, and intersectionality theory.^3^ ^33^ ^34^ Participatory methods such as co-production and co-design show promise as they are particularly valuable to engage marginalised groups.^35^ ^1^ For example, photovoice (PV) can disrupt the traditional relationships between researcher and participant.^36^ ^28^ How an issue is defined or viewed, and by whom, might be key to problem solving.^37–39^ PV has been used to engage service users in co-creation of their healthcare.^39^ ^40^ ^41^ It also provides a safe, supportive, and creative approach to iteratively deepen personal narratives, whilst accommodating variations in literacy, ethnicity, and levels of disability.^36^ ^42^

### Participants and recruitment procedures

Participants were within two years of at least one CAT episode under the powers of the Mental Health Act (England & Wales, 1983, 2007; MHA), and living in one of eight locations in England (Oxford, London, Birmingham, Leeds, Derby, Bradford, Manchester, Lancashire). These venues were chosen to include a mix of urban and rural areas, and variation ethnic group makeup of residents.

Purposive sampling ensured participants were from diverse ethnic groups, across the 8 health systems, the fullest age range (over 18), and representative of gender and specific civil and crisis sections of the MHA (Sections 5(4), 5(2), 4, 2, 3, 136,).^*^ We excluded those who had only experienced forensic sections. The design sought to ensure an ethnically diverse sample overall and not to compare findings by ethnic groups.

Community organisations and NHS trusts assisted with recruitment. We involved potential participants in coffee mornings and by advertising through local charities and the National Service Users Network (NSUN), NGOs, radio channels, as well as through the project’s Public Participant Involvement Research Group (PPIRG) and its networks. We ensured that those under-represented in research were supported to participate, for example, by paying for carers. We took advice from the PPIRG on study design, recruitment, execution of the research, and members were involved in the interpretation of the data.

### Photovoice procedures

The rationale, methods, and full protocol are already published.^46^ Briefly, participants took photographs, reflected upon and captioned them. They were aware this study sought to understand the use of CAT and identify preventive opportunities taking account of their biographies and complex histories. Photovoice workshops (see Table 1) were conducted in person or online to accommodate the challenges around the COVID-19 pandemic. This also optimised recruitment and participation for those with medical problems. In person workshops were conducted in accessible, public spaces, away from NHS settings wherever possible. Padlet was used to complete roadmaps, document captions, and share images for online participants. Participants received a £15 amazon voucher to thank them at the end of each workshop.

**Table 1:**
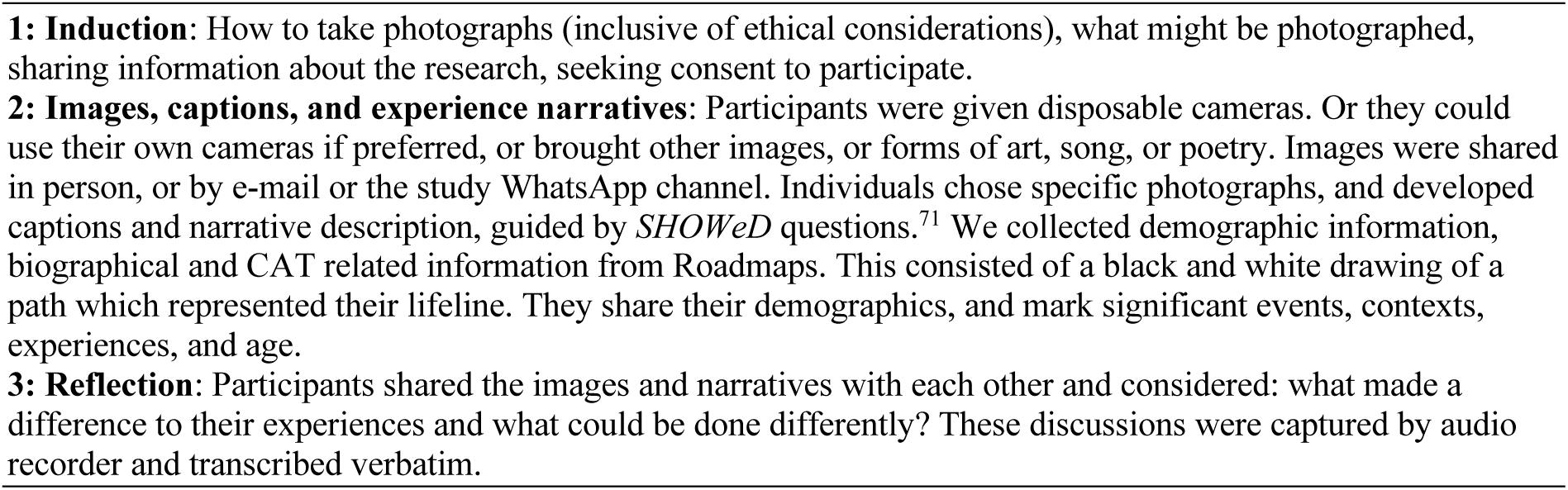
Three Photovoice Workshops.

### Ethics

The study was granted REC and HRA approval [21/SC/0204], and local NHS research department approvals.

### Data Analysis

There are many diverse traditions of undertaking and analysing photovoice.^47^ We have published our approach.^48^ We used *Framework* to organise the data, as it is used for a wide range of qualitative analytic methods,^49^ making the process explicit and transparent, permitting replication, and checks for consistency. The charts were a convenient way to present summary information for interpretation by participants, the PPIRG, and the research team. Images and transcripts were explored using the principles of polytextual thematic analysis^50^ of original images alongside transcripts held in a qualitative software package (MAXQDA). We undertook a descriptive content analysis of images. The photographs acted as an elicitation process allowing participants to gently enter the study, deepen reflections, and explore experiences of CAT at their pace. Then we identified patterns and synthesised the findings using constant comparison.^51^ RM and KB independently coded the information from two research sites, about 40% of the data, to develop and refine a coding framework.

RM coded the remainder of the data, and KB reviewed all transcripts for coding checks on the entire data set. KB undertook the thematic analysis, reviewed by RM.

## Results

Of 60 recruited participants, 48 completed all three workshops. There were three broad types of data: a) demographic and roadmap information for 48 participants, b) 535 images and captions, c) transcripts of 32 hours of discussion (averaging around 90 minutes for online participants, and 2.5-3 hours for in-person workshops).

### Demographics

Most participants were ethnic minorities (see Table 2 for demographics); there was an equal gender balance. The roadmaps demonstrated high levels of multimorbidity (35.2%, 17/48), adverse childhood experiences (45.8%, 22/48), concerns about the impact of their detention on their children, (31.3%, 15/48) and police contact (26.8%, 10/48). Image codes that occurred on more than one occasion are listed in Table 3.

**Table 2:**
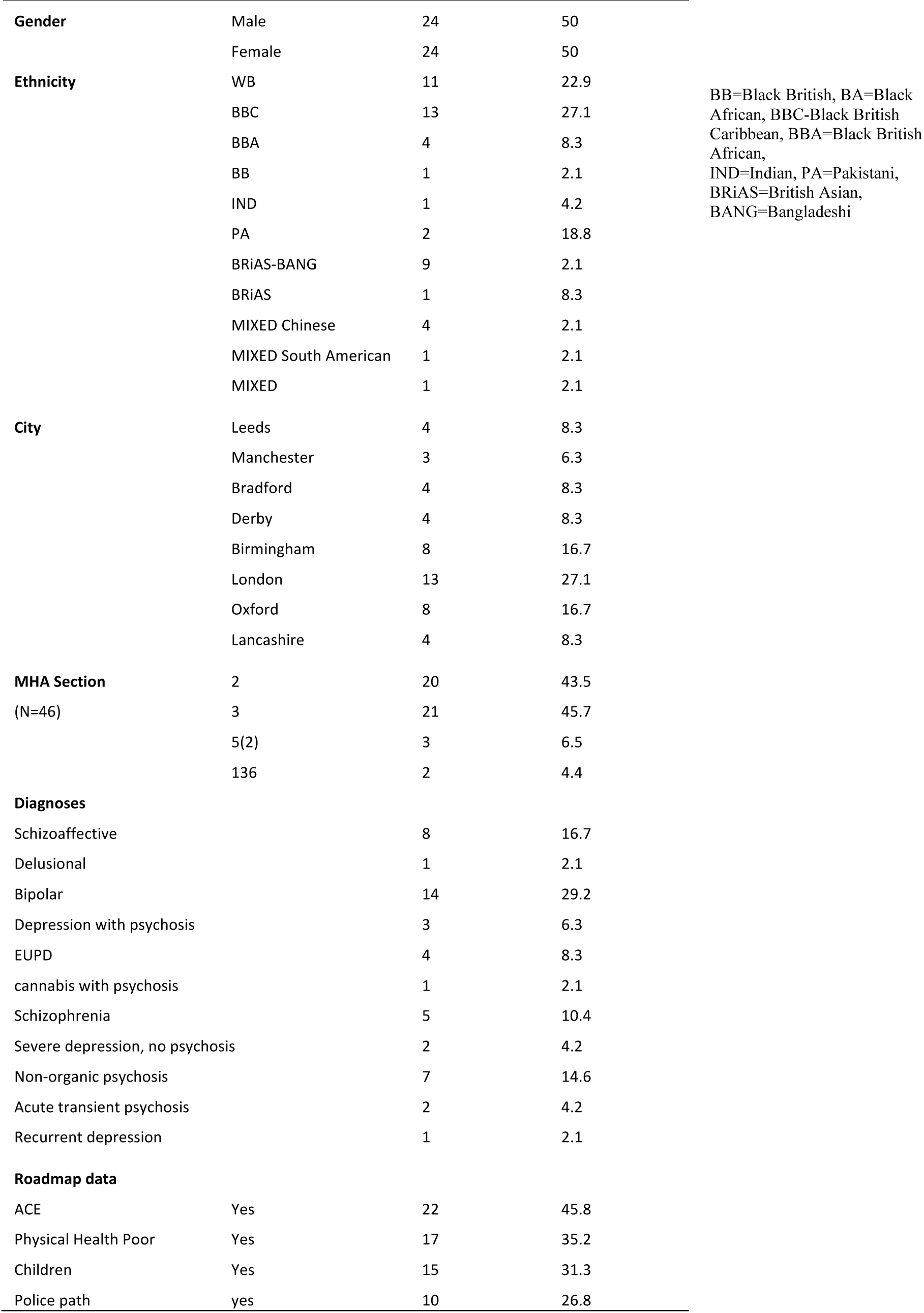
Co-Pact Participant Demographics (N=48, except were different N specified)

**Table 3:** Content analysis of images/captions (mentioned more than once)

|  |  |
| --- | --- |
| Freedom | 2 |
| Cooking | 3 |
| Nature | 4 |
| TV movies | 3 |
| Quote | 2 |
| Water | 2 |
| Clothing | 2 |
| Photo frames | 2 |
| Food/drink | 2 |
| Activities on ward | 2 |
| Sun | 2 |
| Phone | 3 |
| Drugs | 3 |
| Smoking | 4 |
| Door | 3 |
| Lock | 2 |
| Creative tasks | 5 |
| Family | 3 |
| Babies | 2 |
| Trees | 4 |
| Walls | 3 |
| Gates | 4 |
| Medication | 11 |
| Fields gardens | 7 |
| Venues | 2 |
| The participant | 12 |
| People | 7 |
| Flower | 12 |
| Ornaments | 6 |
| House | 9 |
| Windows/blinds | 3 |
| Body parts | 5 |
| Religious symbols | 4 |
| Covid | 2 |
| Community venues | 6 |
| Hobbies | 11 |
| Bed | 3 |
| Events occasions | 2 |
| Remembrance 2 | 2 |
| The past | 4 |
| Family | 6 |
| Music | 6 |
| Cars/vehicles | 5 |
| Pets | 9 |
| Animals | 7 |
| Seaside | 3 |
| Green spaces | 16 |
| Indoors | 17 |
| Outdoor | 12 |

### Thematic Analysis

First, we report an overall inductive thematic analysis and synthesis of the transcripts (see Figure 1; Annex A shows a matrix of quotes for overall themes). Annex B contains images from 6 participants and their captions to illustrate the overarching themes (colour codes and a key signify gender, age, ethnicity, location). The full library of images is online (https://www.flickr.com/photos/198122100@N06/albums/72177720307479644/; https://www.psych.ox.ac.uk/research/chimes/co-pact/photovoice-exhibition). We then report less common but important insights, referring to participant characteristics by aggregate summaries of gender (M,F), ethnicity (in accord with their preferred term of those used in the ethnic coding in the census: White British-WB, Black Caribbean-BC, Black British-BB, Black African-BA, Bangladeshi-BA, Indian-Ind, Pakistani-P, Asian-A, mixed-M), and age-group (decades).

**Fig 1.**
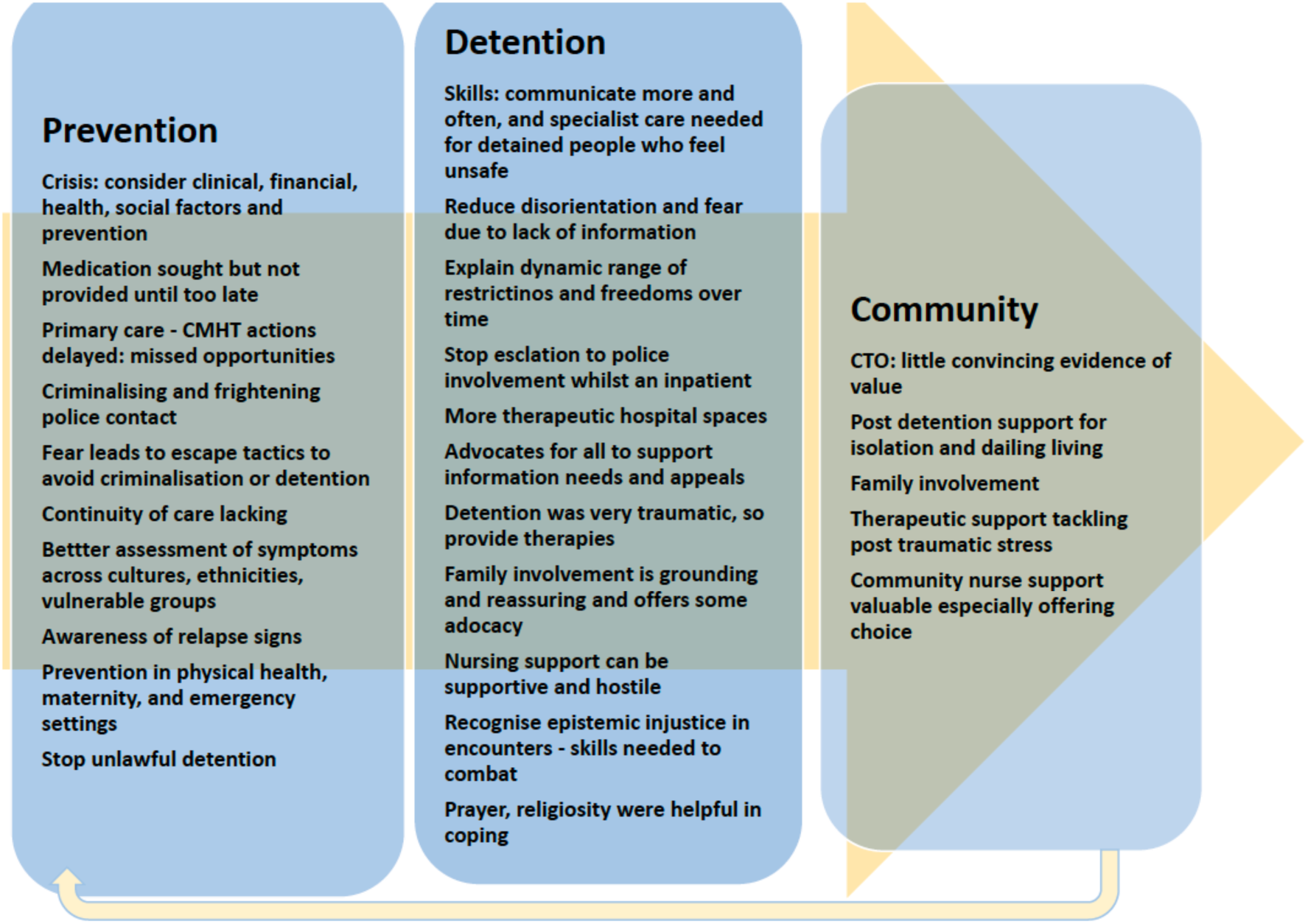
Thematic Analysis of Experienced Data.

### Overarching Findings

#### Awareness, Crisis Pathways & Prevention

Participants were aware of deterioration in mental health and sought intervention early. Despite this, many did not receive any preventive intervention and were admitted in crisis. For example, when a primary care team was contacted, they alerted the community mental health teams (CMHT), which was unable to respond in a timely way. Financial difficulties were sometimes the trigger for a relapse and admission. The participants suggested professionals needed to be more aware of the role of poverty in mental health crises, and financial interventions might avert stress and prevent CAT.

#### Families and Carers

Participants felt that their loved ones were excluded. When present, they were able to offer advocacy, and protection against poor care by demanding accountability of staff. Families were often not permitted to visit or did not visit. Active involvement of family, friends, or carers was considered important to help patients in distress feel calmer. Maintaining these relationships helped restore dignity by reminding people of their lives outside hospital.

#### Detention processes

Some detention decisions were influenced by the availability of beds and not the need for admission. In some sites, people reported travelling for 4-5 hours to be admitted away from their home. Two participants in different parts of the country described being detained and transported in an unmarked van, which felt like ‘being kidnapped’ as there was nothing to indicate these were health service vehicles.

#### Coercive Care and Staff Interactions

There were strongly held views that many mental health professionals appeared to not have sufficient experience, knowledge, or skills to support people in distress at the time of admission. Participants recognised it was challenging to care for very unwell people, and they were appreciative of the demands on staff. However, they felt there was too much variation in the levels of knowledge and skills amongst staff (for example, by day and night shifts), and too much stigmatising language. There were accounts of staff responses being hostile, dehumanising, disrespectful and threatening. Staff were sometimes openly rude and aggressive. Participants felt coerced. They could not understand why some professionals did not follow better professional standards, with care and concern for patients. In one instance, a person described being forced to have medication as ‘like being raped’.

The lack of interaction with professionals was a source of dissatisfaction. Communication was often through small openings in doors. The only interactions appeared to be to give medication or to record observations. Shortages of staff also meant leisure activities were not available, as risk management was prioritised. There was evidence of good practice too.

Activities such as painting or quizzes were very welcomed and found to be helpful especially at weekends, which were otherwise very quiet.

The accuracy of medical records was questioned, as was confidentiality. For example, when secluded and blamed, participants feared the official documentation did not reflect their experiences and the records were inaccurate. Their voice was not allowed to come out, despite being detained for longer than six months in some instances. One participant described a diagnosis being shared with the employer without permission, resulting in long standing unemployment.

There were positive comments, for example, of community nurses acting with compassion and being more flexible than inpatient staff. Specifically, community staff were willing to trial different potentially helpful approaches, whereas inpatient care seemed less flexible or inattentive to needs. Our study took place during the COVID-19 pandemic thus the inpatient restrictions and staffing pressures might have influenced these experiences. Reassuringly, some participants said they trusted staff to give advice and do the right thing, it was good to know they ‘had my back’.

#### Therapeutic Environments

Participants felt waking on a ward with no explanation was disorientating, especially as professionals wore plain clothes. Given their mental state, they did not know they were in a safe place. Participants wanted more information repeated on several occasions, to understand options and learn what was happening to them. They asked that professionals take more time, repeat the messages, use different modalities, for example, in induction packs.

Participants were uncertain what ‘detention’ meant in terms of which freedoms they had at different points of their hospital stay, and options for securing more freedom; they received little information about why seclusion decisions were made, or what was happening when secluded. Once, detained, there were few opportunities for exercise or ordinary things, with no access to families or friends. In some environments, the facilities fell short: no bedding, no toilet seat, no daylight, with nursing staff isolated in their station or office. ‘It felt like a zoo’, and so noisy that they could not sleep. It was not perceived to be a good place to recover, although short stays were tolerable.

### Specific Findings

#### Police Involvement

Thirteen participants commented on police involvement ^(4 women; ethnicity: 4WB, 2P, 5BB, 1BC, 1 mixed ethnicity; age: one aged 10–29, two 20–29, four 30–39, two 40–49, three 50–59, one 60–89)^. Unexpected police arrivals were distressing and embarrassing when seen by neighbours: ‘everyone watched’, ‘they drag you out sometimes with handcuffs and kicking.’ One participant rang the help number provided by the hospital on boxing day, the police arrived with a social worker. He was rude to them at first and only later talked calmly and thanked them. The process of detention was slow, and he knew it could escalate, moving from one section to another. Participants felt powerless and criminalised by police involvement, and sometimes took actions to keep safe in vulnerable moments. One participant ^(M, P, 20–29)^ over expressed symptoms of mental illness when arrested or when the police arrived, saying he ‘played the mental illness card’ to get to hospital and avoid being criminalised. Being handcuffed and removed from their homes was demeaning. Many said hospital admission often required a police arrest first; one person became accustomed to ‘making use of the police’ to ensure admission and avoid a criminal justice pathway.

Participants appreciated the police when they were kind but feared escalation and the use of force. A woman ^(F, BB, 30–39)^ was surrounded by police unexpectedly when on the way home. She had stopped in a park to ‘rest and get some air’. The participant had not realised she was on the train tracks and not a park. In this instance, police intervention was helpful. Participants acknowledged that the police did not enjoy the role and were good at the job: ‘they could be more understanding than paramedics’ ^(F, WB, 30–39)^. Although participants recognised the police fulfilled a valuable role, they complained sometimes they were rude. Another person in a police cell was handcuffed; he took a dislike to the officer and provoked him; the police helped by asking him to take medication, saying he would become a voluntary patient. He was getting agitated and apologised as they handcuffed him. He was in a cell overnight and later diagnosed with a brain tumour as the underlying problem. Another participant ^(M, mixed race, 50–59)^ was on a medical assessment ward for two weeks and was surrounded by elderly men receiving end of life care. This did not help his mental state and so he ran away and jumped in a reservoir; he was suicidal; the police rescued him and returned him to the ward.

Police involvement was sometimes a consequence of the participant’s actions; for example, if a person feared someone was breaking into their house. On arrival of the police, the person feared being kidnapped when asked to go into the back of an ambulance. On refusing the participant was arrested, put into a van, taken to the ward, and placed in an isolated room without explanations ^(M, WB, 10–19).^ The police were called by inpatients when a female patient was being racist towards a participant ^(M, P, 20–29)^, in order to ensure the incident was not overlooked; and when a participant was pushed by another inpatient ^(F, BC, 50–59)^. A senior ward nurse called the police in response to requests for help and medication. The participant was ignored for over a half an hour, whilst seeking medication to calm agitation. The staff member became angry, ignored the participant, even turned her back, and called the police as a deterrent. The participant was then placed in a seclusion room, which was disorientating as there were no windows. She could not tell if it was day or night, and it was cold. Then, she was threatened with being ‘*IM’d’* (intramuscular medication against her will).

This account showed an unnecessary escalation, and the use of power and coercion rather than a therapeutic relationship to negotiate needs. This could easily be dismissed as a patient’s misperception; participants were aware of this possibility. However, participants said dismissal of their views was common. This was recognised as a structural problem, of power and knowledge of professionals over-riding that of the patient.

#### Preventive Actions

There were nine people who referred to prevention ^(4 women; ethnicity: 4 WB, 1 P, 2 BB, 1 CB, 1 mixed ethnicity; age: one 10–19, two 20–29, three 30–39, three 50–59)^_. Participants recognised medication could prevent_ detention. There were examples of professionals encouraging reflection on signs of relapse and plan to prevent detention. A Punjabi speaking doctor discussed the situation with the participants mother in her native tongue. A consultation in a group setting reiterated the importance of medication in prevention. This participant felt the community teams and support were brilliant and she had a good relationship with the community team. She reflected on how they seemed harsh, but it was for her own good: she emerged ‘out of the fog and back to her senses’.

Another participant had been discharged from community mental health services after six years of remission. She recognised an emerging psychosis but could no longer access a crisis team for medication and relapsed over 2 years. The medication was not so helpful in fact, due to adverse effects, but CBT prevented a relapse for quite some time. There were examples of early actions by patients to prevent admission, and they were awareness of relapse signatures. However, it took too long, in one instance, six months from contacting the GP to being seen in specialist services; then a fifteen-minute meeting left the person feeling unknown and misunderstood. This meant the person did not trust the GP’s suggestions. A woman with a postpartum psychosis spoke of services during the pandemic. She had 5-10 minutes with a midwife and all activities were centred on the child’s health. She felt stressed and had a family history. As a single mum, she felt her mental health should have been noticed and relapse prevented. She was separated from her child for a week’s admission.

#### The Dynamics of Communication and Assessment

Contact with a care coordinator was perceived as important and helpful when there was a trusting relationship ^(F, P, 20–29)^. More could be done to prevent admission when psychosis was emerging ^(M, WB, 30–39).^, by sitting with the community or primary care team. This participant still believed the content of her delusional beliefs, but did not talk openly about them, anticipating the negative response. She said sharing information made her seem more unwell to the professional. Another participant ^(F, BC, 20–29)^ asserted that problems started long before hospital admission, and the diagnostic and formulation process was not culturally sensitive or precise, with excessive negative reading of experiences and behaviours. Misdiagnosis and misinterpretation were argued to explain higher admission rates for ethnic minorities.

#### Community Treatment Orders (CTOs)

Only two participants (a Pakistani women and a Black British man, respectively) talked about CTOs. One said CTOs were used when there was a be shortage rather than for any therapeutic reason, although he felt it could prevent relapse. One participant was negative about CTOs and perplexed why a CTO was used if he had never been violent:

‘It felt like an ASBO (Anti-Social Behaviour Order).’

‘…I will be recalled…it’s like prison…you get back to a locked ward. What’s the answer?’

#### Emergency Departments

Seeking help from Emergency Departments was not a good experience ^(F, WB, 30–39)^: ‘you’re pushed aside and looked down on.’ A participant ^(M, mixed race, 50–59)^ who attended the emergency room after self-cutting, said as blood dripped from her arm, she was seen by generic staff not mental health specialists: ‘…at what point are you an emergency…you need help. You don’t have physical scars. How far do you have to go’ [to get help].

Contrary to guidance and legislation, some were held in emergency departments despite not being on a section ^(M, WB, 30–39)^. ‘The security guard was preventing me from leaving…he had no right to, even though I needed to be prevented from leaving…’. There were positive experiences: ‘….security guards, ambulance staff, emergency staff, everybody was lovely’.

#### Feeling Safe and Advocacy

Participants wanted more information at the time of detention. Furthermore, isolation in uncomfortable surroundings reinforced a lack of safety. Participants felt unentitled to comfortable and humane environments when detained; they questioned why such conditions were considered reasonable (police cells, seclusion rooms, and assessment and inpatient facilities). One participant ^(M, 60–69, WB)^ wanted a private room and someone to talk with (mental health staff not emergency staff); he resisted taking medication because he feared being ‘like a zombie’. He suggested staff should instead ask more about feelings as these were not easily expressed. Isolation and loneliness were common experiences, some wanting more communication and a person with them all the time especially in moments of crisis.

_There were mixed experiences of advocates_ ^(3 women; ethnicity: 1 WB, 2 BB, 1 P; age: two 20–29, one 30–39, one 50–59)^, for example, some advocates supported the doctor’s position and insisted the patient take the medication. For example, a person’s mother urged she take medication. Just as she agreed, she was transferred from an assessment section 2 (lasting 28 days) to a section 3 for treatment (lasting up to six months); she felt this had happened because in a ward round she had persistently refused medication^(F, WB, 50–59)^. Some participants felt everyone should have an advocate, as ‘you’re not in a state of mind to decide …when first detained’. One participant found the advocate helpful in explaining rights, getting a solicitor, ‘someone on your side when you’re in there.’ One participant felt the advocate helped end the section by articulating their case, organising evidence, and reassuring the participant that someone cared and doing what they could not manage on their own. It was the advocate who explained what was happening to them rather than the ward staff. ‘You rarely get told your rights, and even if on the ward, not everyone is aware; and people have different capacities to process information’.

Some were unable to secure an advocate until released into the community, too late to help with appeals. The paperwork required for an appeal was thought too burdensome ^(F, WB, 50–59)^, especially when a person was distressed. One participant ^(F, P, 20–29)^ won three of four appeals, with support, the right staff, approach, paperwork and reports. The advocate assisted. This participant was troubled about the ‘life and death’ decision in front of three judges, ‘like you’re stuck in prison…it was a big thing’ and she did not feel prepared. An advocate and more information would help.

#### Detention Trauma

Eight participants ^(five women; ethnicity: 4 WP, 1 Ba, 1 In, 1 BB, 1 P; age: two 20–29, two 30–39, two 40–49, one 50–59, one 60–69)^ said the process of CAT was traumatic (see Table 4). They talked of isolation, loneliness, losing control over everyday decisions, and feeling distressed. Several spoke of ‘being broken’ by the experience rather than seeing it as helpful or restorative. For those with pre-existing post-traumatic symptoms, additional traumatic events were more distressing. One person was unable to remember what had happened until friends explained, as distress and re-traumatisation can affect memory processing. This participant had just taken an overdose and recognised she was not ‘in the right mind’. The cubicles in A&E were ‘small and stressful’ and she ‘wanted to leave and get some air. There were five security guards for one person (small frame) which was stressful. The situation was escalating, then one guard persuaded her to calm, ‘he took a risk and it worked’. She spent 8 hours in the early hours in A&E, eventually a nurse explained she was to be detained.

**Table 4:**
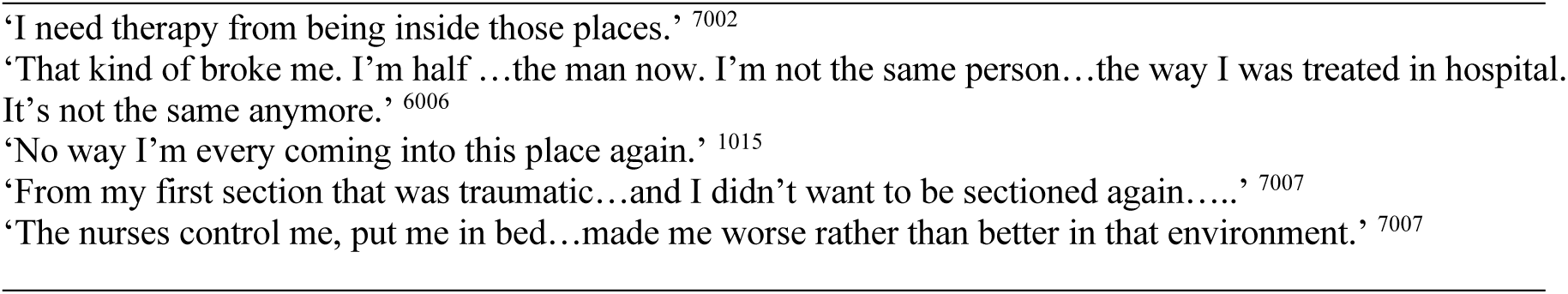
Traumatic Detention.

#### Community Support

Leaving hospital was liberating ^(F, mixed race, 50–59; F, P, 20–29)^, giving access to gyms and shops. However, ‘adjusting back into living on your own in a quiet place^’^ was difficult in contrast to the ‘noise and number of interactions on a ward’. There was a need for structure outside the hospital, by joining health groups in the community; the only formal advice on leaving was around taking medication (F, WB, 30–39). Participants felt being discharged without carers or a network of community meant people became unwell again and returned to hospital (Table 5).

**Table 5:**
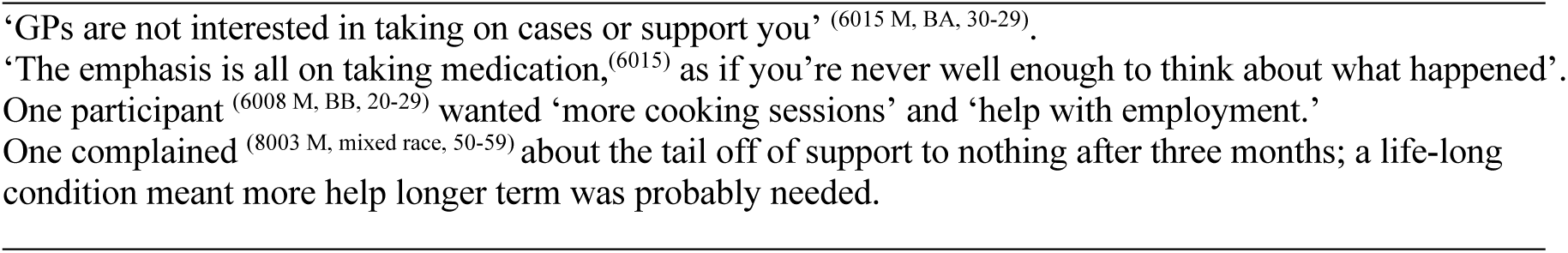
Community Supports.

#### Religion and Ethnicity

Religious practice was helpful, for example, ‘praying when ill’ ^(F, P, 20–29; M, BB, 40–49)^. Yet there was often no place to do so on the ward. A multi-faith room in one site was spoken of very highly as an example of good practice. There were positive experiences of chaplains but also apprehension about evangelical and orthodox sentiments being expressed ^(F, WB, 50–59)^. White members of staff told one participant to ‘believe in God; this was not racist… but uncomfortable.’ One chaplain helpfully spent an hour with the person, but the variation between hospitals and over time was a concern. The participant was a Buddhist, and there was no religion-specific support for her. Another ^(F, British Asian, 30–39)^, a Muslim, was not given choice of appropriate foods.

#### Race and Epistemic Injustice

‘Many minorities don’t understand their rights, they need advocates, and then the language and communication barriers also hinder recovery’ ^(F, P, 20–29)^. One person ^(F, BC, 20–29)^ was admitted whilst ‘grieving for her nan’. She was distressed and trying to get support. She was prescribed antihistamines and then accused of taking alcohol when leaving a message for her Community Psychiatric Nurse. She had not taken alcohol. She ‘broke down completely’ for not being taken seriously and being discredited.^52^ She complained and was dismissed as just grieving, rather than receiving more mental health care which she needed. She also witnessed a patient with a strong Caribbean accent was also dismissed and ignored. A lot of staff misinterpreted her as aggressive. One participant ^(M, BC, 50–59)^ felt black people were admitted more rather being given home treatment.

Multi-ethnic wards created an atmosphere in which people had to hide concerns or conflicts. Participants talked of how (in some wards) the nurses were mostly black; some staff from India would speak only to men, and other staff would refuse to speak English, leading to conflict. Racialised explanations can arise between staff, between patients, and between staff and patients in the absence of skills to talk about racism and discrimination more generally.^3^ ^53^ Victimisation (by staff) of the same person was also witnessed, whilst others were left alone even if they infringed the rules, like smoking on the ward. Some positively noted ^(M, WB, 30–39)^ that there were staff of many ethnicities, and they were caring; whereas other participants (see above) found this to be a source of tension and potential conflict. Yet, having staff who looked like the patient was helpful ^(F, BA, 20–29)^.

#### Logic Model: designing preventive interventions

Taking all the findings, we developed a logic model showing the context, inputs, processes, outputs, and impact (see Figure 2). This summarises the elements of interventions which our participants felt would substantially change their experiences and outcomes, anticipating touch points to co-design future interventions in later stages of the research.

## Discussion

This study brings experience to the fore, through images and text, sharing with us the struggles and repeated systemic obstacles to recovery.

### Epistemic Injustice

Epistemic injustice includes testimonial injustice,^54^ the discrediting of information on the basis of demographics and in this instance due to ethnicity, diagnoses of mental illness, and loss of agency due to CAT. Participants communicated many instances of hostility and discrediting of their views, not being given appropriate information, and nor having clarity on choices or appeals processes. This happened before admission and during. In part, cognitive impairments in the process of psychosis may make some people vulnerable to poorer function and need for more intensive care and support, which may end in relapse if not provided.^55^ Rather than respond to requests for care, for example, with more and repeated information, they were ignored or the system failed to respond adequately, leading to relapse and ultimately CAT; or escalation to seclusion and police involvement whilst an inpatient.

**Fig 1b.**
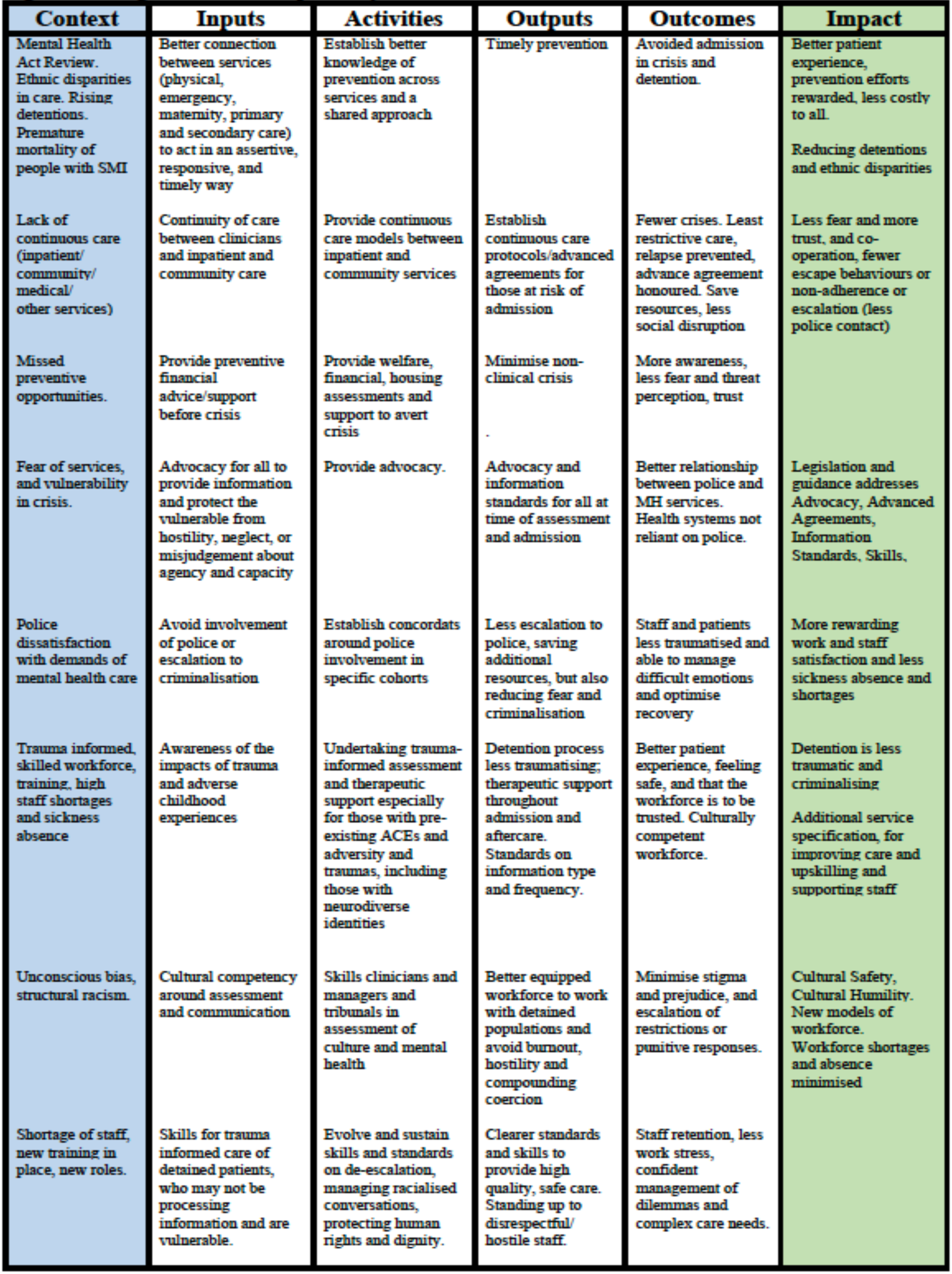
Logic Model of Proposed Systems Interventions.

These cycles of discrediting, disempowerment, and escalation to coercive care could and should be prevented. The main complaint was a failure of earlier care and the quality of care once detained. Shared decision making can be useful,^56^ but needs explicit skills, and standards and competencies suitable for people receiving CAT. Bridging community, inpatient, general medical, social care and even specialist care like maternity, all require new models of identifying those at risk of CAT and commensurate assessment and decision-making skills. Integrated care systems and calls for more generalists rather than specialists irrespective of profession might help tackle both stigma and more cohesive care systems for prevention. Epistemic injustice also includes hermeneutics, how our structures of knowledge production, providing public services, and solving problems may themselves be perpetuating inequalities.^54^ Advocacy and advanced agreements can assist,^54^ as can progressive participatory research methods in knowledge production as these contest concepts such as mental illness, diagnosis, and disparities in care that reflect of historical relationships between patients and statutory agencies.^52^ Thus services need to reflect more complex models and consider the intersections of racism, migration, religion, and complex trauma, alongside age and gender, rather than only crude ethnic group or diagnostic classifications.^28^

### Prevention

We found scope for more preventive work before a crisis, and better resourced services equipped for preventive strategies. These should bridge and encompass primary care, specialist mental health care, maternal care, emergency departments, and other social supports. There is scope for financial and social interventions among those facing hardship and adversity. Importantly, these are not financial or social incentives for behaviour change, rather they seek to reduce distress that leads to crisis.^57^ The data were not strong or consistent on CTOs as few participants commented. We need alternatives to admission and detention, and long-term investment, better professional skills and standards to provide greater service capacity, flexibility, supported by clearer inter-sector policies.^22^

### Police Involvement

Police involvement in the mental health system is hotly debated,^58^ ^59^ some arguing the police are essential to manage crises and risk. Others argue the police should never be involved, and only if there are criminal activities. Police involvement was felt to be stigmatising and criminalising, provoking fear of escalation, perhaps adding to the avoidance of care options and open resistance. The experience of detention can be felt as deeply racialised, and impossible to separate from racism in society.^60^ ^61^ Police involvement already leads to criminalisation of racialised groups, and criminal justice pathways to mental health care have been questioned for some time.^1^ ^62^ Efforts to reduce police involvement are controversial, but may prevent policing-related escalations of crises.^63^ This requires services and professionals to acquire appropriate skills and to shift these long standing traditions.

#### Therapeutic Environments

Worryingly, therapeutic, respectful, and safe care were lacking, further compounding fear and a lack of trust. Supporting people experiencing traumatic symptoms before, during and after CAT, assuring family involvement in all stages, and personalisation of care to support religious practice were recommended for future service design. Many participants had experienced childhood traumas making CAT even more distressing. Family therapy, CBT, and trauma-informed therapies require development, testing, as this may help prevent readmission and future CAT episodes.^64^ ^65^ Although racial trauma is recognised,^66^ the impact of detention as trauma is important to further evaluate, not least given the potential links with criminalisation and fears of state authority and harms, something that is a historical reality that people fear today.^67^

#### Strengths and Limitations

We developed a logic model for preventive interventions and reducing CAT. We developed novel methods of participatory research using photovoice and demonstrated an approach to collating and analysing image and narrative data. The approach permitted diverse racial and ethnic groups and lived experience experts to engage when previously they would not have entered research.^52^ They felt like equal partners in generating and interpreting new knowledge for wider impact.^§^ The peer researchers and lived experience experts were central to all stages of the study, including communications, problem solving, analysis, and preparation and dissemination of outputs. Lived experience experts reviewed the data and interpretations, and drafts of this publication.

Although we experienced recruitment challenges, such as accessibility of complete ethnicity and section data, our data set of 60, of which 48 produced narratives and images across three workshops, is one of the largest on this topic according to an excellent meta-synthesis;^68^ this meta-synthesis reported 17 of 56 studies included ethnicity; only 2 had comparable sample sizes or slightly larger to ours (up to 59); of these two, one was a review and included 20 people of ethnic minority status; another recruited 49 patients of which only 8% were ethnic minorities with detention experiences. None of the reviewed studies were seeking to specifically target and underserved group that does not usually participate in research.

Recruitment through screening routine data appeared to not be sufficient to engage potential

#### Conclusions

These lived experiences data show many missed opportunities for prevention attuned to the needs of those being detained. We demonstrated testimonial and hermeneutic forms of epistemic injustice, to promote the development and implementation of more effective interventions. Our findings are consistent with previous studies which emphasise staff skills, de-escalation, collaborative care and the integration of services are effective at reducing detention.^69^ Our findings provide new mechanisms of Self-Determination Theory through psychological safety, therapeutic environments, pervasive contexts, and life experiences.^70^

## Supporting information

Annex A & B

## Data Availability

The data set includes pictures and captions which is publically available.
There are also transcripts and extracts, which are not public to prevent patient identification. However, synthesised data and anonymous extractions can be made available if appropriate ethical approval are considered for secondary analysis. This is a qualitative data set so requires specialist skills in secondary data analysis.

## Acknowledgements

This study/project is funded by the NIHR-PRP (NIHR201704). The views expressed are those of the author(s) and not necessarily those of the NIHR or the Department of Health and Social Care.

Conflicts of Interest: none declared

**Research in Context**

### Evidence before the study

Escalating rates of compulsory admission and treatment (CAT) and racial and ethnic disparities in CAT are well established in the UK and European and North American countries. The reasons for these findings are debated, and often attributed to individual illness influences, rather than structural problems in services. People from racial and ethnic groups are under-represented in research, and so their voices do not enter policy or practice. Yet, legislative and policy and practice reforms require better lived experience data, including those from racial and ethnic groups.

### Added value of this study

We developed a logic model of how coercive care and CAT could be prevented by co-design of future interventions to address the preventive opportunities exposed in experience narratives. We found epistemic justice in care processes, a lack of preventive actions in response to help seeking, dismissal of preventive efforts by patients, excessive use of police and coercive care, and insufficient attention to culturally and racially and religiously competent assessment and care. New skills standards are needed for people receiving CAT, including trauma-informed inpatient and aftercare, new standards for levels and methods of providing information during crises, and more skills on de-escalation when receiving CAT. Advocacy and advances agreements could be incorporated into trauma informed care and discharge planning.

### Implications of the study

The findings expose epistemic and structural mechanisms through which people receive CAT followed by coercive and unhelpful care. We set out preventive opportunities, and make recommendations for both practice and policy reforms, as well as for new legislation, specifically in England and Wales, but the findings are instructive for care sectors more widely. We also show new ways of undertaking inclusive research and set out the methods for peer research and lived experience to be fully incorporated in all aspects of study design and delivery, with implications for commissioners of research.

## Footnotes

* Sample size in qualitative research is driven by study purpose, depth of data collected, and volume of data, rather than minimum levels as is common in statistical studies. Although samples of 12 may be sufficient for saturation among homogenous populations,43. Vasileiou K, Barnett J, Thorpe S, et al. Characterising and justifying sample size sufficiency in interview-based studies: systematic analysis of qualitative health research over a 15-year period. *BMC Medical Research Methodology* 2018;18(1):148. doi: 10.1186/s12874-018-0594-7 we emphasized notions of <u>information power</u> (the more information the sample holds, relevant for the actual study, the lower number of participants is needed)44. Malterud K, Siersma VD, Guassora AD. Sample Size in Qualitative Interview Studies: Guided by Information Power. *Qualitative health research* 2016;26(13):1753-60. doi: 10.1177/1049732315617444 [published Online First: 2015/11/29] and <u>adequacy</u> determined final sample sizes.45. Hennink M, Kaiser BN. Sample sizes for saturation in qualitative research: A systematic review of empirical tests. *Social Science & Medicine* 2022;292:114523. doi: https://doi.org/10.1016/j.socscimed.2021.114523 Thus, we were careful in assessing the sampling as we proceeded.

§ We will be archiving all images for online access, creating case studies, and touring an exhibition for the public, policy makers and specialist. participants, as it was not grounded in trusting relationships. However, during the post COVID recovery period, there were staff shortages and pressures on clinical time. Some services enforced COVID policies that prevented community-based recruitment and data collection.

