## Supplementary material for "Racialised experience of detention under the Mental Health Act: a photovoice investigation of practice, policy and legislation": Annex A & B

#### ANNEX A: Overall Thematic Matrix

| Overall Themes Matrix | Mental Health Awareness | Prevention | Detention process | Coercive Care | Staff interactions | Detention as trauma | Ward Activities | Community Care | Family involvement |
|---|---|---|---|---|---|---|---|---|---|
| **Sense of self** | "I learned so many new words to the point where it's actually very bizarre how many new words I've had to learn in order to keep up with what's being said and what I found is that" (male, Asian British Indian, 21-30) | "Yeah, because my care coordinator had said that part of the research was to reduce the amount of people that get sectioned and get seen it in the homes instead...but I found that personally getting sectioned was beneficial to me. It actually helped me. So that's from my point of view, it was necessary." (male, Pakistani, 41-50). | "It's a tough one, though, because I mean, I don't know how well I was to even be making judgments for myself or have, I don't know..." (male, Chinese-British, 31-40)<br><br>"I think advocates is a big one as well, because when you get sectioned in hospital you get asked if you want help from an advocate, but when you are in that state of mind you don't really think properly do you, so I think everyone should just get given an advocate, automatically." (2001, Manchester, F2F, female, Pakistani, 21-30) | "And then there was a time where they had to inject me because I was too kind of chaotic within the ward itself, and I screamed "Rape." That's how, that's how intensive it was, because I felt as though they were going to go ahead and rape me at the time of the treatment. No one said to me that we're giving you the injection." (female, British Asian, 31-40). | "it makes you feel like not, like you're worthless human being who has got no opinions of their own, no rights, no feelings, no nothing, it makes you feel worthless, it really does just because you know you need help and support because you are deteriorating or because you are mentally unstable or something like that it doesn't mean you are not a human, we are still humans you know what I mean, we should be treated with a bit of respect at least, not oh you're ill so you know, ah it really gets me angry" | "That kind of broke me. I mean, I'm half of the man I am now. So, it's kind of why I am. I'm not the same person. So the way I was treated there in hospital, it's it's it's not the same anymore. " (male, Bangladeshi, 31-40)<br><br>"And I feel like I need to get closure on it or talk about it to understand what happened like it is almost a trauma just being in there, isn't it?" (female, WB, 31-40). | "We did a lot of things with them, we did colouring, painting, which was really therapeutic. In addition to that, we did things like quizzes which were really really good. The OTs used to do a quiz, every...I think it was two during the week and it was really interesting."<br><br>"Although that's quite a nice colouring book but the ones in hospital were... I mean I just thought, "I'm not a 3 year old, I really don't want to do this" (female, WB, 51-60). | "I found that my community psychiatrist was a lot more understanding than the hospitals' ones, he's happy for me to try different medications or try come off them with his support" | "my two sisters have been really helpful. When I did get ill initially, they saw the signs and got me in touch with a psychiatrist." |
| **Communication** | "where the doctor wrote that thing on my sicknote. And that is something that I can neve ever forget. Because I think it effects me a lot. Because I'm not, I'm not encouraged to find another job. Because that is always in the back of my mind. What if they ask why did you stop working here? " (female, Black British Caribbean, 31-40)<br><br>"Putting me under that bracket like, it's like tell an employer that he's got schizophrenia, it's like wow. That's my perception, straight away., it's like stigma." | "you need to really start looking for information about what it is you have a diagnosed with it, and how can you just work your way forward to be sane or healthy again or whatever you want to call it? Because it's not like you're being a horse with a disease that it's not going to be taking away from you forever. So it's either you can manage it or you can cure it. " (male, Black British Caribbean, 21-30) | "am I allowed to go to town today you know I need to get a few bits? Oh no you're on a section 3, I'm on a section what? I just lost it then I thought what am I here for 6 months now? Its not nice that" (F2F, female, Pakistani, 21-30)<br><br>"He started saying: "well, yeah, we're going to, you know, you're not well enough yet." She did that once to me...but when I was feeling better to be able to say: "look, I want to have leave." You know, because I was more confident and I was able to answer their questions." (female, South Asian British, 61-70)<br><br>"They kept saying I won't get sectioned, they trick you into taking you into hospital and that's what messes with your head," | "if you're under section you have no option, you are on drugs." (male, mixed race, 51-60).<br><br>"you witness something and you your voice doesn't come. You are nothing. So it's only one. I'm in the right course. You know, if the records they write, imagine someone attack me or something or vice versa. You know, the records are wrong. It's going to be wrong forever. This is only one side of the story." (male, Black British African, 31-40). | "apart from the healthcare workers, the staff nurses don't really have much interaction with you apart from giving you medication and to tell you off for doing something wrong" (female, Black British Caribbean, 51-60).<br><br>"I found care, compassion with one of the nurses who was very kind and came across with me and heard me on the same level, rather than "I'm a nurse and you're a patient and I don't really care", we felt quite close. (male, Pakistani, 41-50). | "that's the seclusion room, they just put you in there like you're to blame, like this is your fault; They don't explain anything to you, it's like you're stripped of all your rights." | "actually being able to focus on something other than your own mental health, that was really impossible to distract yourself." (male, mixed race, 51-60). | " I know someone has got my back out in the community like I've had her for a year now and she is the one who told me about this so, that helps in the community having a care co-ordinator someone of your own you can turn to for help" | "So like he never, ever got contacted before my ward round. And I just think that's really bad because it's his life as well, like and he's just as important as me." |
| **Environment** | "There's not enough ads out there, there's not enough...how do you say it? There's not enough stuff on the tv. There's not enough..." (female, WB, 31-40 ) | "There's a lot of factors that effect mental health. A lot of factors, like running out of money for me is one of the factors that maybe can start, trigger the fact that I won't be sleeping right. " | " communication, like some posters or something to say where you are, what's happening to you. Just that you're in a hospital to start with because it's not clear you're in a hospital at all. You, you wake up and there's plain-clothed people in your room standing over you. And everybody's plain clothed and you don't, you don't- how can you realize you're in a hospital being treated when everybody's just dressed in their own clothes, looking the same as everybody else. And the walls are painted with children's things, pictures, really confusing. And there's no way to say that you're where you are and that you're in a hospital at all". (female, WB, 31-40) | "ward please believe me, they are like zoos. (laughs) When you go in there, you're like: wow: You know everything is just wow. (laughs). All day people are banging on the glass init. It used to drive me mad. Yeah, for the office. Because in there the office is locked. It's not like on other units where they are open, the office is locked up there. For people, bang for attention. Do this, do that. And it's all day" (male, Black British Caribbean, 31-40). | "I feel like ward rooms really highlight the power imbalance between service users and service providers. Again I feel like it should be mandatory that there is an advocate or a third party who isn't connected to the NHS at all: (female, Black British, 21-30). | "There's no bedding, no toilet seats, I couldn't see outside, I could just see the staff observing me from the glass windows. It felt like a prison cell, especially when they would communicate with you through the flap in the door. " | "the bench meant a lot to me actually being allowed out...being for a section and not allowed out at all and then eventually just let out for 15 minute walks and things supervised and eventually unsupervised."<br><br>. | "that's the thing about that type of hospital, it's not the place to recover properly anyway and you get out and get out into the community and that's better for care" | "I'm just lucky that my mum and my dad, they were just so supportive while I was in there, not one day went by where they didn't come see me, not one day, they always brought in food for me. I was eating good food" (male, Asian British Indian, 21-30).<br><br>"if my partner and my family could visit there more it would have been an easier ride" |
| **Service constraints** | | "This very last time I went in, I phoned my GP about a week or so before, but they referred me to like the community mental health team, and the referral took too long. So I didn't actually see anyone before I got so unwell that I had to go in." | "The last time I was sectioned, I was picked up in a black van, it was three people in black clothes, I was completely... I didn't know was going on and they go so fast all the way down,? Something like that, I didn't have a clue what was going on I thought I was being kidnapped, it was really scary" (female, WB, 31-40).<br><br>" I did have a mental health assessment and I think they actually, obviously not the sole reason, but I think a lot of the reason they didn't section me was because they literally didn't have anywhere to put me." (female, WB, 31-40) | "I personally feel that they're not trained or equipped for it because they're like "How do I treat this person? How do I handle this person when they're going through an episode or they're having a psychosis or a breakout?" You know, so the first thing they think is "sedate, restrain, inject," but really in theory, it could be, you know, help them with a breathing technique or, you know, help them to calm down or give them some water or take them into a cool, air conditioned room." (male Black British Caribbean, 21-30). | " I understand that if a place is short-staffed, then probably the first thing to go is going to be a painting activity rather than a one-on-one observations." (male, WB, 31-40) | | "When you are first detained you are not allowed to go out you know, it depends on the section you are on. It's awful in it, tell you the truth. One minute you are exercising, then you can't exercise and you can't eat the food you eat. So you have to kind of like start readjusting." (male, Black British, 31-40). | | "And then after a couple of days I was there, I just couldn't sleep and of course, because of COVID couldn't have any visitors" |
| **System Challenges** | "I was going to say, you said about discrimination about like race, obviously I don't, well, not obviously, but I don't have experience of that, but I did have experience of like feeling discriminated against because of my diagnosis, even in like mental health settings. I feel like people see what I'm diagnosed with and then like make assumptions.' | In reference to crisis centres "I would appreciate those. I would appreciate those because those are the times you don't want your family" | "Yeah I just want NHS, I just want to know at what point you go to A and E and they consider you an emergency. you need help. You don't have physical scars, you're feet not dropping off. At what point. How far do you have to go. " (female, Black British-Caribbean, 31-40).<br><br>"Because they'd normally, the psychologist or whatever. They always say you are not unwell, crisis café or take the, go to A and E. But when you go to A and E, they do blood tests, they do pressure check, the regular vitals. " | " I was just asking about how the whole medication: what's in the medication and stuff like that. And the nurse in charge just told me to just shut up and just take it. So, I feel like, you don't really have a voice in some hospital settings because of the people in charge, their characters, and how they perceive you as a patient, that you just do as you're told and you have no right to question them." (Black British African, 31-40). | "Some mental health staff are really poorly trained in my opinion...the requirement level of taking people into the into the work place is really really basic at the moment and is really, really concerning because they take in anyone that coming from anywhere because the staff levels aren't really, really concerned about, about how these people are trained and how they perceive when the first line to get the job screening" (female, South American British, 31-40). | "also sometimes there'd be six of us patients and about 20 staff. And you think, there's supposed to be a shortage? How come you, you know, they'll be standing in a group chatting and ignoring us! " | "There's a, for example, there's a gym, but you weren't allowed to use the gym, you know. So there was there wasn't much that a patient could actually do while on the assessment ward just to feel a little bit like, you know, you could- except maybe if you were lucky enough to find a book that you wanted to read, " (male, mixed race, 51-60). | "I think CTOs are, for me it is, its working out for me the CTO because the shortage of beds as well like you can't get emergency beds, say I've deteriorated, and I need help right now, the CTO being in place its helping init I can get a bed if I need to there" | "I think if you don't, if you're psychotic and you don't see your family, it just delays the recovery. Like you start to realize things that you're thinking and believe in are more real when you see your family and you speak to your family." |

Annex B

### Images from six people illustrating general themes

Tall, thin, standing still just like your emotions.

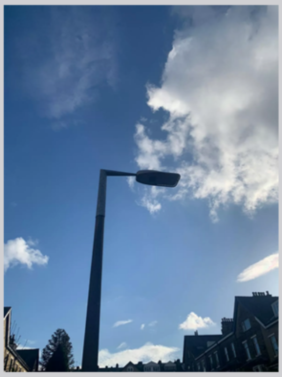

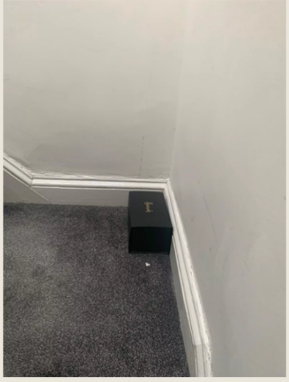

In hospital your stuck in a corner of the room in hospital ward with no where to go. Just like being trapped inside this box in the corner of the room.

Where it leads in both directions in hospital weather you are well or not.

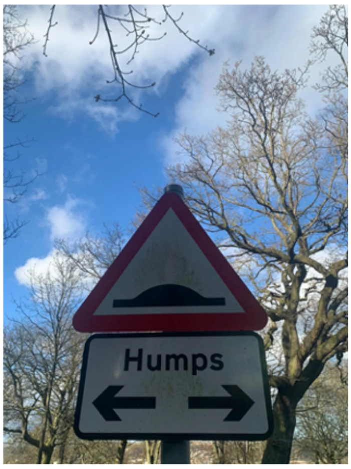


That's where the medication should go as some of it does not help at all.

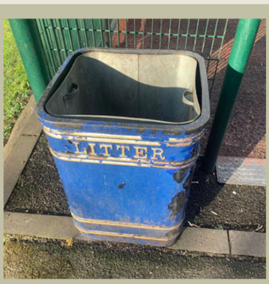


I took this photo to represent being discharged. Trying to convince everyone that I was better. Reflecting on the 'event', going back to 'normality' and freedom.

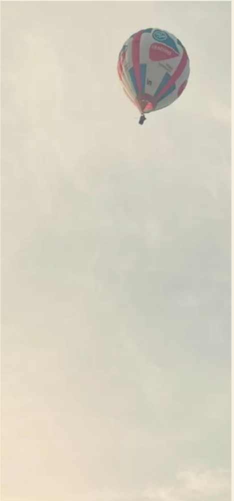

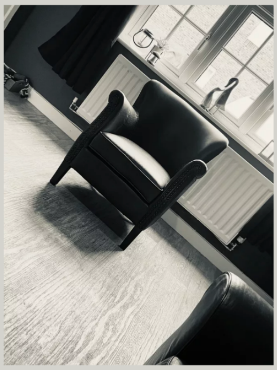

I took that because the last two times I was sectioned, the doctors and the AHMP came to my house. They sat in that chair. This chair represents impending doom. I was having to explain something I didn't understand and it felt like an interrogation.

This shows missing home. Wondering if they are all ok. Have I upset them? Are they coping? They need me but I can't help them.

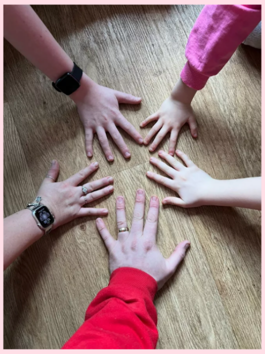

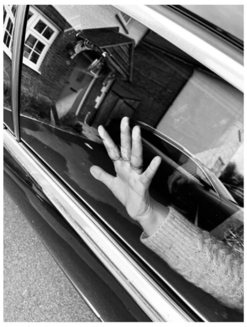

This symbolises the journey of being taken to hospital. The first time I was sectioned I was taken to hospital in a black van by three people not wearing uniforms. I thought I had been kidnapped. It was scary, confusing and lonely. It's like a journey into the unknown.

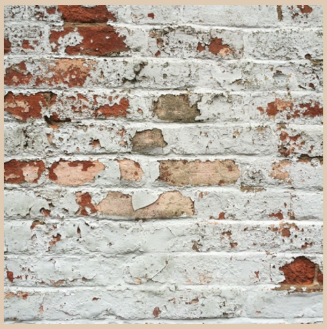

Banging your head against a brick wall…
Sometimes I find mental health care/accessing mental health care can be like hitting your head against a brick wall. Being detained is all well and good… as long as there's a bed for you. Which often there isn't. I've been on the 'bed list' for 5 weeks now (for informal admission this time). Multiple weekly incidents, police and ambulance involvement… and still…. 'No beds'.

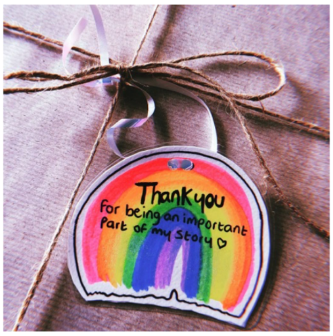


Evidence. The evidence of a head banging episode whilst detained. Blood on the wall. This is as bad as it gets.

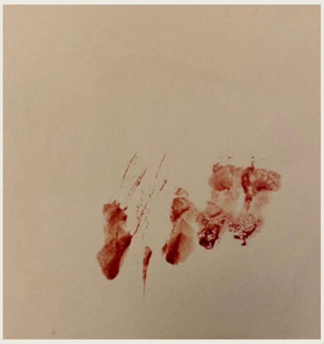

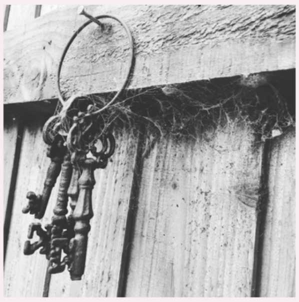

Lock me away and throw away the key. It's normal to you, you get people admitted everyday…. But to me it might be my first time away from home. It's scary. It's lonely. It feels like the end of the world. Please try and remember that.

The people are everything. You can get a really posh building, high tech with brand new sofas and TVs and en suites. But what REALLY matters, is the people. The nurses, the HCAs, the students, the doctors and other ward staff.

Instead of being detained in hospital its better to see the Home Treatment Team.
I prefer that because I'm still in the community.
Being detained is the same as prison because you're not allowed to go nowhere.

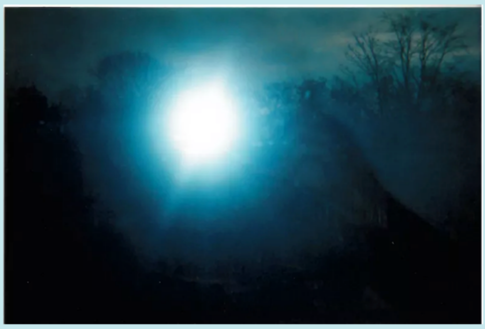

I took this picture because I like the view of it. It had just snowed. It was quiet, no one around. Fresh air.

I was going on the train from Floweryfield to Guidebridge. Going to meet one of my friends.

Going to see a friend I first met on the ward when I was first sectioned (2014/2015).

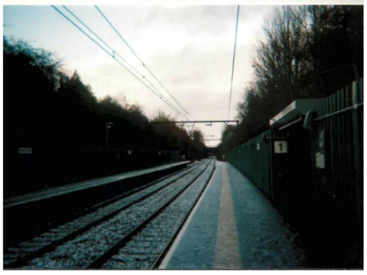

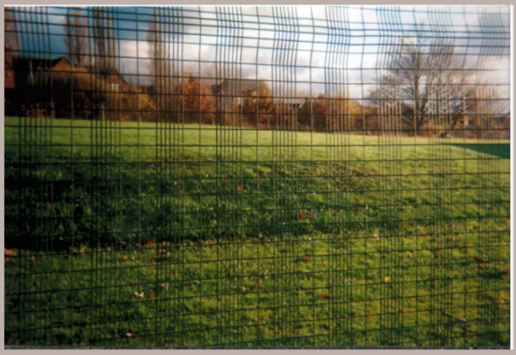

This picture is about exercise.
It's good to exercise every day.
Good for your mental health.
But when I was detained I could not exercise for a while.
It a good thing to exercise but under detention is impossible.

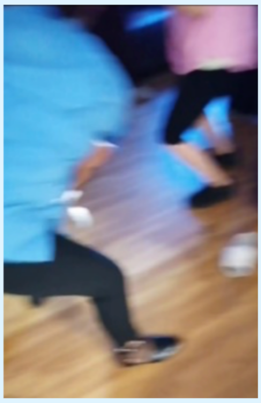

This was such a fun day on the ward. It made a difference to the fighting, shouting and general discourse on the ward that myself and others found distressing. One of the patients was listening to music and getting her hair weaved by a HCA. I came over and started dancing. Then sooner or later HCAs and patients were joining in on the dancing fun in the lounge. I've never seen anything like it nor have I heard anything like it. Having a moment like that to bring us all together makes a world of difference and seeing that the HCAs helped weave someone's hair really touched me. I don't think people realise how meaningful those small gestures are when you're on the ward and stripped of a lot of your outside comforts.

There needs to be a better system to highlight ways to seek advocacy and makes complaints while on the ward. It was frustrating trying to be taken seriously a lot of the time. I remember seeing a consultant when I was on PICU a few days after I was brought out of seclusion. I was starting to piece things together about my reasons for being sectioned and I mentioned that I think I had a first episode of psychosis. He laughed and said "people who are going through psychosis don't realise they are going through psychosis". I had to retort that I was training as a peer support weeks before my admission, in hopes of joining the early intervention in psychosis team at an under 25s mental health service. He just responded with "ah, I see . . .". I felt like my voice didn't matter in moments like that. Ward rounds are very unnerving. Having support in place would have helped me deal with the ward rounds better.

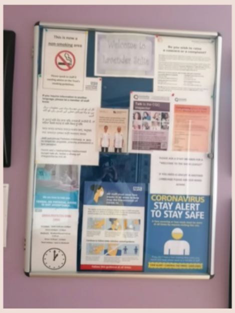


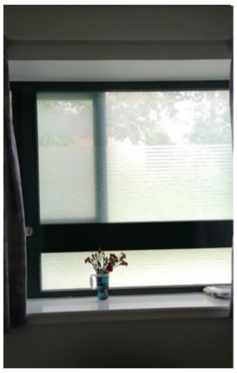

Even though I still wasn't comfortable about the lack of privacy on the ward, this room on the acute ward was more comfortable than the rooms I stayed in at the PICU. I felt like a prisoner on PICU the first time I came to my senses when I was in an anti tear outfit in the seclusion room.

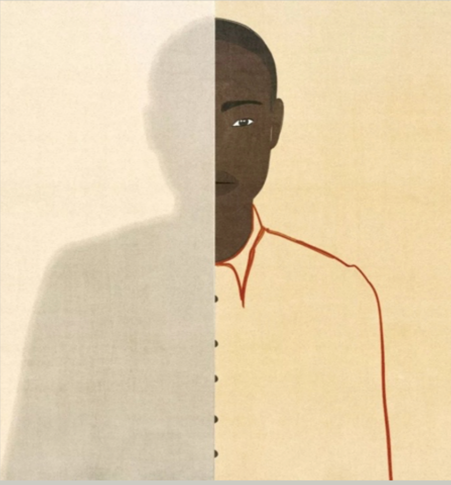

There is a part of myself that remains unknown to me. Little gaps in my personality as a result of thoughts and experiences which I have no control of. My thoughts have their own voice.

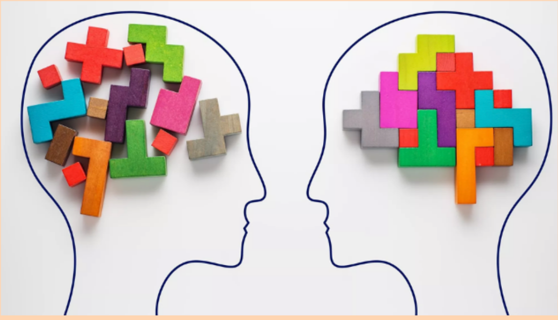

This is how it feels when my brain starts to play games on me and then the medication fixes it.

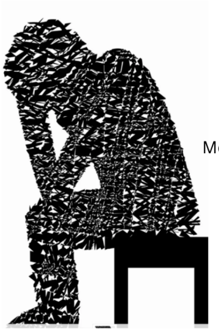

Mental self inflicted Bullying.
